# Educating the Next Generation of Cancer Researchers: Evaluation of A Cancer Research Partnership Training Program

**DOI:** 10.1101/2023.05.15.23289973

**Authors:** Lin Zhu, SJ Dodd, Yuku Chen, Emily R. Kaminsky, Zhiqing Elaine Liu, Grace X. Ma, Olorunseun O. Ogunwobi, Carolyn Y. Fang

## Abstract

African American, American Indian and Alaska Native, Hispanic (or Latinx), Native Hawaiian, and other Pacific Islander groups are underrepresented in the biomedical workforce, which is one of the barriers to addressing cancer disparities among minority populations. The creation of a more inclusive biomedical workforce dedicated to reducing the burden of cancer health disparities requires structured, mentored research and cancer-related research exposure during the earlier stages of training. The Summer Cancer Research Institute (SCRI), a multicomponent 8-week intensive summer program funded under the Partnership between a Minority Serving Institute and a National Institutes of Health-designated Comprehensive Cancer Center. This study assessed whether students who participated in the SCRI Program report greater knowledge and interest in pursuing careers in cancer-related fields than their counterparts who did not participate in SCRI. Successes, challenges, and solutions in providing training in cancer and cancer health disparities research to improve diversity in the biomedical fields were also discussed.

## Introduction

### Background

The underrepresentation of minorities in basic and clinical research is a barrier to addressing cancer disparities among minority populations [1–5]. African American, American Indian and Alaska Native, Hispanic (or Latinx), Native Hawaiian, and other Pacific Islander groups are underrepresented among those who earn a Bachelor of Science degree, doctorate degree, and among those in the biomedical workforce [6]. In 2019, underrepresented minorities (URMs) were awarded 11.7% of science and engineering research doctorates, while comprising approximately a third of the US population. Additionally, URMs with science, engineering, and health doctorates held 8.9% of academic positions, which is considerably lower than their share of the population [7]. Enhancing early mentorship of emerging scientists from URM communities has been shown to increase exposure, awareness, and preparedness for graduate studies in the biomedical field and may reduce the attrition observed at later stages of the academic pipeline [8–12]. Data from recent studies suggest that undergraduate cancer research experiences are effective in increasing interest in cancer research and enrollment in a graduate or professional school [13, 14]. For example, one study evaluating an undergraduate program designed to increase the representation of URM students in oncological research found that 69% of the participants reported graduate or professional school enrollment, with 45% having completed an oncological program. Participants in this study also expressed that working with a mentor motivated them to continue on to a career in research [14]. Together, these studies suggest that the creation of a more inclusive biomedical workforce dedicated to reducing the burden of cancer health disparities requires structured, mentored research and cancer-related research exposure during the earlier stages of training.

The Synergistic Partnership for Enhancing Equity in Cancer Health (SPEECH) is a comprehensive regional cancer health disparity partnership between Temple University/Fox Chase Cancer Center (TUFCCC) and Hunter College (HC), funded by the Comprehensive Partnerships to Enhance Cancer Health Equity (CPACHE) U54 grant mechanism of the National Cancer Institute [15]. The purpose of SPEECH is to reduce cancer health disparities among underserved minority populations in the Pennsylvania-New Jersey-New York City region through cancer disparities research, community outreach, and career development for underrepresented early-stage investigators (ESIs) and students [15]. The Partnership is comprised of 5 Cores including the Administrative Core (AC), the Research and Education Core (REC), the Planning and Evaluation Core (PEC), the Bioinformatics and Biostatistics Information Core (BBC), and the Community Outreach Core (COC). The main goal of the Research Education Core (REC) is to support educational activities that enhance the training and mentorship of a diverse workforce to meet the nation’s cancer research needs. The Core capitalizes on the numerous strengths in training and research education across TUFCCC and HC to facilitate the professional enrichment of students and ESIs underrepresented in cancer research. A key component of the training supported by the REC is the Summer Cancer Research Institute (SCRI), a multicomponent 8-week intensive summer program based at Temple University/Fox Chase Cancer Center, which serves between 10 and 17 students each summer. The express aim of the SCRI is to provide cancer research education and training opportunities for students, especially those who are from underrepresented minority backgrounds, and to increase the medical and research pipeline of people focused on addressing cancer health disparities.

The primary objective of this study was to evaluate whether students who participated in the SCRI Program report greater knowledge and interest in pursuing careers in cancer-related fields than their counterparts who did not participate in SCRI. In addition, this study examined the successes and challenges as well as solutions in providing training in cancer and cancer health disparities research to improve diversity in the biomedical fields.

### Program overview

The SCRI Program is held each summer on the TUFCCC campuses. In the SCRI program, students participate in hands-on research training in laboratories under the mentorship of established investigators. In addition, students participate in a didactic curriculum that includes cancer seminars, skill-building workshops, journal clubs, social activities, and a research symposium at the conclusion of the 8-week program. A mentored, cancer-focused research project is the cornerstone experience.

### Recruitment

Eligible SCRI trainees must be currently matriculated at Temple University or Hunter College as an Undergraduate or early (first- or second-year) graduate student. Information about the SCRI Program was broadly disseminated across both campuses through targeted multi-channel strategies, including flyers, emails, classroom visits, and social media posts. Informational sessions with SCRI mentors, alumni, and REC leaders were also held to promote the program and engage interested students.

### Application and selection

All applicants submitted a formal application in which students shared their research interests and experiences and described how the program may improve their academic/professional development. The application included an optional item that allowed applicants to describe or explain any challenges they may have encountered during their academic career. All essays/responses were capped at a 500-word limit. Along with the essays, applicants were asked to submit a resume/CV and an unofficial academic transcript. Applicants had the option of submitting up to two letters of recommendation, but this was not a required component of the application.

During the competitive application process, all submissions were reviewed by a committee of 25 to 30 reviewers. The review committee consisted of REC members, previous-SCRI mentors, SCRI alumni, SPEECH partnership members, and early-stage investigators. Each application was reviewed by at least two individual members. Reviewers scored each applicant based on GPA/academic background, research interest/experience, and communication/writing skills. Then, using the NIH 9-point scale (with ‘1’ representing an exceptional application and ‘9’ representing a weak application), reviewers assigned an ‘Overall Rating’ for each applicant. The top third of highest-ranked applications were discussed by the review panel during a group conference call, where reviewers conferred about each candidate’s strengths and fit for the program, and then selected the finalists to invite into the SCRI Program.

## Data & methods

### Participants and procedures

To evaluate the impacts of the SCRI program on participants’ knowledge on cancer and cancer health disparities and their interest in pursuing higher degrees or careers in related fields, as well as their satisfaction with the SCRI program, we conducted a cross-sectional survey with accepted SCRI applicants (i.e., participants) and their peers whose applications to the SCRI were not accepted (non-participants) from four cohorts (2019 – 2022). Email invitations with a link to a REDCap survey were sent to all 51 SCRI participants and 488 non-participants with the goal of identifying any differences between the two groups in knowledge and career goals. In total, 32 SCRI participants and 47 non-participants responded to the survey from June to July 2022. Data from one SCRI participant was excluded from the analysis due to extensive missing data, thus yielding 31 SCRI participants. Printed versions of the surveys are provided in a supplementary file to this article. The Temple University Internal Review Board reviewed and approved this project (protocol number 29481). Written consent forms were obtained from all survey participants. Only the lead author (LZ) had access to information that could identify individual participants during and after data collection.

### Measures

We measured respondents’ knowledge in three domains: (1) cancer health disparities, (2) cancer biology, and (3) cancer prevention. For each domain, we used four questions to assess knowledge (Supplement A). Participants received 1 point for each correct answer. We calculated knowledge scores by summing the points from all questions in each domain. Examples of questions from each domain included, “Compared to non-Hispanic white women, how likely are African American women to die of colorectal cancer? (a. more likely, b. just as likely, c. less likely, d. don’t know)”, “Which of the following is the current “gold standard” for evaluating the efficacy of novel cancer treatments? (a. animal studies, b. phase 1 clinical trials, c. phase 2 clinical trials, d. phase 3 clinical trials, e. case-control studies)”, “What is the recommended screening test for lung cancer? (a. sigmoidoscopy, b. blood test, c. low-dose computed tomography (LDCT), d. Papanicolaou test).” The knowledge score for each domain ranged from 0 to 4, with a higher numeric value indicating a higher level of knowledge. We then computed a total knowledge score by summing the three sub-scores. The total score ranged from 0 to 12.

Respondents’ interests in pursuing higher degrees or careers in a cancer-related field was also measured. Examples of questions in this domain included “How interested are you in pursuing a career path in cancer biology or cancer health disparities research in academia?” and “How interested are you in pursuing a career path in cancer biology or cancer health disparities in the industry?” Response options were: “not interested at all”, “somewhat interested” and “very interested.” For subsequent analyses, we combined the first two categories.

### Analytical approach

Descriptive statistics were used to characterize the demographic features of the two groups (see Table 1). We conducted t-tests to examine potential differences between SCRI participants and non-participants on knowledge scores of cancer health disparities, cancer biology, and cancer prevention, and their level of interest in pursuing higher degrees or careers in cancer or a cancer health disparities related field. Among SCRI participants only, we examined their level of satisfaction with the program. The survey data were accessed for research purposes and analyzed between November 2022 and March 2023.

**Table 1.** Demographic Characteristics of the SCRI Participants and Applicants.

| N (%) | Participants (N = 31) | Non-participants (N = 47) |
| --- | --- | --- |
| **SCRI Cohort or Application Year** |  |  |
| 2019 | 4 (12.90%) | 3 (6.52%) |
| 2020 | 5 (16.13%) | 8 (17.39%) |
| 2021 | 12 (38.71%) | 16 (34.78%) |
| 2022 | 10 (32.26%) | 19 (41.30%) |
| **Gender** |  |  |
| Cisgender male | 8 (25.81%) | 11 (23.40%) |
| Cisgender female | 21 (70.97%) | 32 (68.09%) |
| Transgender female | 0 | 1 (2.13%) |
| Non-binary, gender fluid, or gender queer | 1 (3.23%) | 2 (4.26%) |
| Prefer not to answer | 0 | 1 (2.13%) |
| **Race** |  |  |
| White | 9 (29.03%) | 15 (31.91%) |
| Black/African American | 4 (12.90%) | 7 (14.89%) |
| Asian | 10 (32.26%) | 17 (36.17%) |
| American Indian or Alaska Native | 0 | 3 (6.38%) |
| Multi-racial or other | 3 (9.68%) | 2 (4.26%) |
| Prefer not to answer | 5 (16.13%) | 3 (6.38%) |
| **Ethnicity** |  |  |
| Hispanic | 5 (16.13%) | 4 (8.51%) |
| Non-Hispanic | 23 (80.65%) | 41 (87.23%) |
| Prefer not to answer | 1 (3.23%) | 2 (4.26%) |
| **First-Generation College Student** |  |  |
| Yes | 17 (54.84%) | 16 (34.04%) |
| No | 14 (45.16%) | 27 (57.45%) |
| Prefer not to answer | 0 | 4 (8.51%) |
| **Academic Level at Time of Survey** |  |  |
| Undergraduate student | 14 (45.16%) | 32 (68.09%) |
| Working in healthcare, medicine, or science having completed a bachelor's degree | 4 (12.90%) | 3 (6.38%) |
| Master's level graduate student | 4 (12.90%) | 4 (8.51%) |
| Working in healthcare, medicine, or science, having completed a master's degree | 4 (12.90%) | 0 |
| Doctoral level student, including PhD, medical/dental student | 3 (9.68%) | 6 (12.77%) |
| Working in academia, having completed PhD and/or MD degree | 1 (3.23%) | 0 |
| Other | 1 (3.23%) | 2 (4.26%) |
| **Major†** |  |  |
| Public health | 5 (16.13%) | 7 (14.89%) |
| Epidemiology | 4 (12.90%) | 4 (8.51%) |
| Nutrition | 4 (12.90%) | 1 (2.13%) |
| Molecular biology | 4 (12.90%) | 3 (6.38%) |
| Cell Biology | 6 (19.35%) | 6 (12.77%) |
| Genetics | 3 (9.68%) | 5 (10.64%) |
| Biochemistry | 5 (16.13%) | 8 (17.02%) |
| Neuroscience | 2 (6.45%) | 7 (14.89%) |
| Psychology | 3 (9.68%) | 2 (4.26%) |
| Medicine | 9 (29.03%) | 18 (38.30%) |
| Nursing | 0 | 6 (12.77%) |
| Engineering | 1 (3.23%) | 1 (2.13%) |
| Computer science | 1 (3.23%) | 1 (2.13%) |
| Teaching/education | 0 | 1 (2.13%) |
| Clinical practice | 1 (3.23%) | 2 (4.26%) |
| Clinical research | 3 (9.68%) | 4 (8.51%) |
| Other major | 2 (6.45%) | 6 (12.77%) |

Note: †Column percentages add up to over 100% because respondents could select up to two majors

## Results

Table 1 presents the demographic and academic characteristics of the two groups of respondents. Over two-thirds of the respondents were from the 2021 and 2022 cohorts (70.97% of the participants and 76.08% of the non-participants). About 70% of the participants identified as Asian, Black/African American, or multi-racial. Of the participants, 16.13% identified as Hispanic and 54.84% were first-generation college students, much higher than the proportion of the non-participants (8.51% as Hispanic and 34.04% as first-generation college students).

With respect to knowledge, SCRI participants had significantly higher scores on cancer health disparities (3.45 vs. 2.26, p = 0.0004), cancer biology (2.52 vs. 1.55, p = 0.0001), and cancer prevention (3.55 vs. 3.02, p = 0.02), as well as the total score (9.52 vs. 6.83, p < 0.0001) than did the non-participants (see Table 2).

**Table 2.** Comparison of Knowledge Scores between Participants and Non-Participants of the SCRI Program.

|  | Participants (N = 31)<br>Mean (SD) | Non-participants<br>(N = 47)<br>Mean (SD) | p-value |
| --- | --- | --- | --- |
| Knowledge of Cancer Health Disparities, range: 0-4 | 3.45 (0.99) | 2.26 (1.62) | 0.0004 |
| Knowledge of Cancer Biology, range: 0-4 | 2.52 (0.93) | 1.55 (1.02) | 0.0001 |
| Knowledge of Cancer Prevention, range: 0-4 | 3.55 (0.68) | 3.02 (1.09) | 0.02 |
| Total Score for Cancer Knowledge, range: 0-12 | 9.52 (1.77) | 6.83 (2.78) | < 0.0001 |

The SCRI participants had slightly higher levels of interest in pursuing a graduate-level degree in cancer biology or a cancer health disparities-related discipline (41.94% reporting “very interested”) and in pursuing a career path in cancer biology or cancer health disparities in the industry (41.94% reporting “very interested”), than did the non-participants (36.17% and 36.17%, respectively), but the differences were not statistically significant (p > 0.05). The two groups had similar rates of pursuing a career in cancer biology, cancer health disparities research, or teaching in academia (see Table 3).

**Table 3.** Comparison of Interest in Pursuing a Career in Cancer Related Fields.

| N (%) | Participants<br>(N = 31) | Non-participants<br>(N = 47) | Total (N = 78)<br>N (%) | Chi-Square<br>(df), p-value |
| --- | --- | --- | --- | --- |
| Pursuing a higher degree (master’s or doctorate)<br>in cancer biology or cancer health disparities<br>disciplines |  |  |  | 0.26 (1), 0.61 |
| Very interested | 13 (41.94%) | 17 (36.17%) | 30 (38.46%) |  |
| Not interested/somewhat interested | 18 (58.06%) | 30 (63.83%) | 48 (61.54%) |  |
| Pursuing a career path in cancer biology or cancer<br>health disparities research in academia |  |  |  | 0.03 (1), 0.88 |
| Very interested | 10 (32.26%) | 16 (34.04%) | 26 (33.33%) |  |
| Not interested/somewhat interested | 21 (67.74%) | 31 (65.96%) | 52 (66.67%) |  |
| Pursuing a career path in teaching cancer biology or cancer health disparities research |  |  |  | 0.02 (1), 0.90 |
| Very interested | 11 (35.48%) | 16 (34.04%) | 27 (34.62%) |  |
| Not interested/somewhat interested | 20 (64.52%) | 31 (65.96%) | 51 (65.38%) |  |
| Pursuing a career path in cancer biology or cancer health disparities in the industry |  |  |  | 0.26 (1), 0.61 |
| Very interested | 13 (41.94%) | 17 (36.17%) | 30 (38.46%) |  |
| Not interested/somewhat interested | 18 (58.06%) | 30 (63.83%) | 48 (61.54%) |  |

In addition, we found high levels of satisfaction with the program among the participants, with over 90% reporting that they “agreed” or “strongly agreed” that the SCRI training experience had a positive influence on their plans for continued education (90.32%), increased skills in cancer or cancer health disparities research (93.55%), increased skills on writing a scientific manuscript (93.55%), increased skills on presenting at a scientific conference (100%), and had a positive influence on their future career plans (90.32%) (see Table 4).

**Table 4.** Satisfaction with SCRI Program among 31 SCRI Participants

| Table 4. Satisfaction with SCRI Program among 31 SCRI Participants |  |  |
| --- | --- | --- |
| N (%) | Strongly agree or agree | Neutral, disagree, or strongly disagree |
| “My research experience through the Summary Cancer Research Institute (SCRI) has had a positive influence on my plans for my continued education.” | 28 (90.32%) | 3 (9.68%) |
| “My SCRI research experience has increased my skills on cancer or cancer health disparities research.” | 29 (93.55%) | 2 (6.45%) |
| “My SCRI research experience has increased my skills to write a scientific manuscript.” | 29 (93.55%) | 2 (6.45%) |
| “My SCRI research experience has increased my skills to present at a scientific conference.” | 31 (100%) | 0 |
| “My SCRI research experience has had a positive influence on my plans for my future career.” | 28 (90.32%) | 3 (9.68%) |

## Conclusions & discussion

The SCRI program was designed to provide training opportunities to URM students. The program took measures in the recruitment and admission stages to ensure that relevant information was accessible to URM students. We found that the SCRI participant and applicant groups had similar demographic profiles, with two exceptions. The SCRI participant group had a higher proportion of individuals who identified as Hispanic (16.13%) than the applicant group (8.51%). The former also had a higher proportion of first-generation college students (54.94%) than the latter (34.04%).

Our findings indicate the potential for an intensive eight-week training institute to increase students’ capacity in cancer research, especially related to their knowledge of cancer health disparities, cancer biology, and cancer prevention. The results showed that SCRI participants had a significantly higher level of knowledge on cancer health disparities, cancer biology, and cancer prevention than that of SCRI applicants. Satisfaction was also high among participants, reflecting an overall positive influence of the program on students’ perceived scientific skills and future career plans.

Interestingly, the results did not indicate a significant difference between groups on motivation for and interest in a career in cancer research. Indeed, it is positive to note that both groups are highly motivated and interested. In fact, since all respondents had applied to participate in the SCRI Program, this likely indicates that interest and motivation related to cancer research was already high in both groups. Promisingly, the results indicate that not being selected into the SCRI also did not deter them from aspirations regarding cancer research. Although, it is also possible that only those who remained motivated and interested responded to our survey and those whose interest had waned were less to respond.

Several limitations of the present study should be acknowledged when interpreting these results. First, the sample does not represent everyone who applied to or was accepted into the SCRI program. As such, findings cannot be generalized to a larger population than the respondent pool. In addition, it is possible that only those who were highly motivated by or highly aggrieved with the SCRI program may have responded to the survey, potentially skewing results and limiting neutral response. For example, only non-SCRI participants who are still interested in cancer health disparities and who are still wanting to be engaged with the U54 Partnership may have responded. Similarly, even though the survey was anonymous, SCRI attendees responding to the survey may have felt pressure to respond in a way that supported the goals of the program. Despite these limitations, the findings offer important information related to the potential for an intensive training program to have a positive impact on URM researchers’ skills and career aspirations.

This evaluation of the Summer Cancer Research Institute sponsored by the Synergistic Partnership for Enhancing Equity in Cancer Health between Temple University/Fox Chase Cancer Center and Hunter College provided positive feedback about the potential for an intensive program targeted at under-represented minority students to enhance their knowledge of cancer health disparities, biology, and prevention. Given that the underrepresentation of minorities in basic and clinical research is a barrier to addressing cancer disparities among minority populations [1–5], findings from this training institute suggest that progress can be made towards diversifying the cancer workforce and support cancer-related career trajectories. In addition, similar intensive training programs can also enhance interest in cancer research specifically designed to address health disparities. Tailoring the program in this way to meet the needs of all participants including underrepresented minority trainees can lead to positive experiences, satisfaction, and most importantly increased cancer research knowledge and relevant professional skills.

## Data Availability

All relevant data are within the manuscript and its Supporting Information files.

## Data Availability Statement

All underlying data is available within the manuscript and its Supporting Information files.

